# Geospatial modeling of pre-intervention prevalence of *Onchocerca volvulus* infection in Ethiopia as an aid to onchocerciasis elimination

**DOI:** 10.1101/2022.01.10.22269016

**Authors:** Himal Shrestha, Karen McCulloch, Shannon M Hedtke, Warwick N Grant

## Abstract

**Background:** Onchocerciasis is a neglected tropical and filarial disease transmitted by the bites of blackflies, causing blindness and severe skin lesions. The change in focus for onchocerciasis management from control to elimination requires thorough mapping of pre-control endemicity to identify areas requiring interventions and to monitor progress. *Onchocerca volvulus* infection prevalence in sub-Saharan Africa is spatially continuous and heterogeneous, and highly endemic areas may contribute to transmission in areas of low endemicity or vice-versa. Ethiopia is one such onchocerciasis-endemic country with heterogeneous *O. volvulus* infection prevalence, and many districts are still unmapped despite their potential for *O. volvulus* infection transmission.

**Methodology/Principle findings:** A Bayesian geostatistical model was fitted for retrospective pre-intervention nodule prevalence data collected from 916 unique sites and 35,077 people across Ethiopia. We used multiple environmental, socio-demographic, and climate variables to estimate the pre-intervention prevalence of *O. volvulus* infection across Ethiopia and to explore their relationship with prevalence. Prevalence was high in southern and northwestern Ethiopia and low in Ethiopia’s central and eastern parts. Distance to the nearest river (-0.015, 95% BCI: -0.025 – -0.005), precipitation seasonality (-0.017, 95% BCI: -0.032 – -0.001), and flow accumulation (-0.042, 95% BCI: -0.07 – -0.019) were negatively associated with *O. volvulus* infection prevalence, while soil moisture (0.0216, 95% BCI: 0.014 – 0.03) was positively associated.

**Conclusions/Significance:** Infection distribution was correlated with habitat suitability for vector breeding and associated biting behavior. The modeled pre-intervention prevalence can be used as a guide for determining priority for intervention in regions of Ethiopia that are currently unmapped, most of which have comparatively low infection prevalence.

**Author’s summary:** Areas with unknown onchocerciasis endemicity may pose a threat to the goal of eliminating transmission: they may re-introduce onchocerciasis to areas where interventions have been successful. Additionally, because the vectors (and thus *Onchocerca volvulus* transmission) have specific ecological requirements for growth and development, changes in these ecological factors due to human activities (deforestation, modification of river flows by dam construction, climate change) might change patterns of parasite transmission and endemicity. To estimate the impact of these environmental changes, we must first identify ecological factors that determine transmission and prevalence. We have employed Bayesian geostatistical modeling to create a nationwide *O. volvulus* infection prevalence map for Ethiopia based on pre-intervention nodule prevalence and have explored the effect of environmental variables on *O. volvulus* infection prevalence. We have also identified areas that need additional data to increase the prediction accuracy of the map. We found that hydrological variables such as distance to the nearest river, precipitation seasonality, soil moisture, and flow accumulation are associated significantly with *O. volvulus* infection prevalence. We show that the pre-intervention prevalence can be estimated based on the ecological data and that predicted prevalence can be used as a guide to prioritize pre-intervention mapping.

## Introduction

Mapping infection prevalence is fundamental for control and elimination because it is used to estimate the disease burden and to design and monitor the impacts of interventions. Often the prevalence data are present in the form of point data at different locations and time points, and are aggregated at different administrative levels (1). However, disease risk is a spatially continuous phenomenon that extends across and beyond administrative borders (2). In addition, mapping strategies change depending on the intended endpoint of the intervention (3): when elimination of transmission is the goal, the spatial heterogeneity in disease prevalence has to be quantified accurately so that appropriate interventions can be implemented and, where possible, implementation and monitoring can be informed by the spatial distribution of infection rather than simply along local administrative organizational boundaries. When resources or accessibility to an endemic region are limited, as is the case for many neglected tropical diseases, such thorough data collection may not be possible and methods to extrapolate likely prevalence would be useful.

Using geostatistical modeling techniques, point prevalence data can be transformed into a continuous spatial prevalence map of varying endemicity (2, 4), rather than reporting binary categorization of areas as endemic or non-endemic (5). These continuous maps can extrapolate the prevalence measures to previously unmapped regions based on the spatial autocorrelation between the prevalence measures and the influence of known ecological and socio-demographic factors. In addition, geostatistical models provide unbiased quantification of the uncertainty associated with the prevalence estimates.

Onchocerciasis is a neglected tropical disease caused by infection with a filarial nematode, *Onchocerca volvulus*, that is transmitted by the bites of blackflies (*Simulium* spp.). The vectors have a specific ecological niche: they breed around fast-flowing rivers, requiring high aeration and oxygen content for larval development (6). The flies show diurnal activity and bite humans living in communities near these rivers (7-9). If the biting blackfly carries the infective stage of the parasite (the 3^rd^ stage larvae, or iL3), the larva leaves the blackfly and enters the human host. Inside the human body, the larva develops into an adult worm and forms a nodule, generally localized subcutaneously. People living with onchocerciasis show a range of chronic clinical manifestations, including onchodermatitis, severe itching, rashes, and visual impairment that may culminate in blindness (10). More recently, it has also been linked with epilepsy and nodding syndrome in children (11, 12).

Onchocerciasis is currently targeted for elimination via community-directed mass drug administration with ivermectin (MDAi), either annually, semi-annually, or, in some areas, up to four times a year (13). *Onchocerca volvulus* infection prevalence is measured using counts of microfilariae (mf) in a small of skin biopsy (skin snipping), physical examination for the presence of nodules (nodule palpitation), or antibody tests that detect the presence of antibodies against the parasite Ov16 antigen (14). Rapid Epidemiological Mapping of Onchocerciasis (REMO) uses nodule palpation in combination with geographic information system mapping, and was used by the African Programme for Onchocerciasis Control (APOC) to map prevalence in twenty countries from 1996 to 2012 (15, 16). REMO revealed that the prevalence of *O. volvulus* infection was patchy and heterogenous across Africa (17) and identified areas for ivermectin intervention (3, 15) using a threshold for treatment set at a nodule prevalence of 20%. Onchocerciasis-endemic communities were divided into hypoendemic (nodule prevalence: < 20%), meso-endemic (nodule prevalence: 20–45%), and hyperendemic (nodule prevalence: > 45%) (17, 18) based on nodule prevalence. However, there are still many areas that are unmapped and in which the infection prevalence is not known (19).

In onchocerciasis-endemic Ethiopia, mapping of prevalence has been focused on the western districts based on the high incidence of onchocerciasis and because environmental factors favor blackfly breeding in these regions (20). In contrast, eastern Ethiopia has been assumed to be free of *O. volvulus* infection, which has generally proven true (21). However, a recent continent-level mapping (19) found that most of the implementation units that were predicted to be suitable for onchocerciasis in Ethiopia were not mapped, posing a risk to elimination goals. In addition, there is high spatial variability of onchocerciasis endemicity in Ethiopia, ranging from 0% in some areas to as high as 84% in some areas of southwest Ethiopia (21, 22).

MDAi started in some Ethiopian hyperendemic foci in 2002 (20) and, to our knowledge, there has not been coordinated vector control in Ethiopia. The shift to onchocerciasis elimination officially began in 2013 with a goal to eliminate transmission by 2020 (20, 22): the program moved from annual to biannual treatment strategy in all the known endemic areas and scaled up treatment to other additional endemic areas which were not treated previously (22). Cross-border coordination of MDAi between transmission foci in northwestern Ethiopia and bordering Sudan is ongoing (13). In some cases, transmission decline without intervention has also been reported (23) but onchocerciasis persists in some areas despite MDAi for a variety of reasons, including challenges with treatment compliance (24-26), civil unrest (27, 28), and lately the COVID-19 pandemic (29). In addition, there has been variation in the history and the frequency of MDAi. Most of the hyper- and mesoendemic districts have been treated over two decades, while in hypoendemic districts, MDAi started around 2014 following the policy shift from control to elimination (22).

There is no national-level baseline endemicity map of *O. volvulus* infection prevalence for Ethiopia, which has created difficulty in quantifying the effect of MDAi on a national scale. Baseline/pre-control endemicity is an important indicator of morbidity and a predictor for the time required for elimination (18, 30-32). In addition, the prevalence measures before intervention provide an unbiased relationship between the infection prevalence and environmental variables. Prevalence and onchocerciasis suitability mapping for Ethiopia in previous studies (17, 19) have been done as part of continental-scale research, although Zouré et al.(17) did not consider environmental factors, and Cromwell et al. (19) used presence-absence data which do not capture the magnitude of the prevalence. Although these studies helped to place *O. volvulus* infection prevalence or risk in a broader ecological and epidemiological context, we have focused on a spatial scale which offers us greater flexibility to explore ecological patterns unique to Ethiopia by incorporating both the magnitude of prevalence and associated ecological variables (33). We develop a geostatistical model for the distribution of pre-intervention nodule prevalence of *O. volvulus* infection prevalence in Ethiopia using this approach that considers spatial variation in environmental and socio-demographic variables. Furthermore, we identify the most important environmental and socio-demographic variables contributing to *O. volvulus* infection prevalence, and present estimates of uncertainty in the predicted prevalence that can be used to target areas for further mapping efforts.

## Methods

### Prevalence data

*Onchocerca volvulus* infection prevalence data with site-specific coordinates for Ethiopia were obtained from the publicly available Expanded Special Project for Elimination of Neglected Tropical Diseases (ESPEN) database (34). Nodule prevalence data were collected as part of REMO mapping before the initiation of MDAi, from the year 2001 to 2012, examining the presence of palpable onchocercal nodules in 30 to 50 adults randomly selected from each surveyed village (15, 20). The protocols for the REMO assessment are available in the published guidelines (35). There were 927 geopositioned coordinates for nodule prevalence in 36,010 people. Any observations from the same geographic coordinates at different times were aggregated by adding the number of cases observed and the number of total tests done before calculating the prevalence.

Although the database contains both nodule prevalence data and skin mf prevalence data, in this analysis, only nodule prevalence data were considered for the geospatial analysis because of the limited number of skin mf sampled (n = 126) before MDAi, which limits its utility for identifying associations between prevalence and environmental variables. That said, skin mf data were used to assess the correlation between skin mf as a measure of *O. volvulus* infection and the nodule prevalence measure, which revealed two outlier observations with very high nodule prevalence but with low skin mf prevalence. These were excluded from the dataset because the low skin mf prevalence could not be attributed to a reasonable cause, such as MDAi, as the data were collected before ivermectin distribution. Thus, the final dataset contained nodule prevalence data from 916 unique sites and 35,077 people (Fig 1).

**Fig 1.**
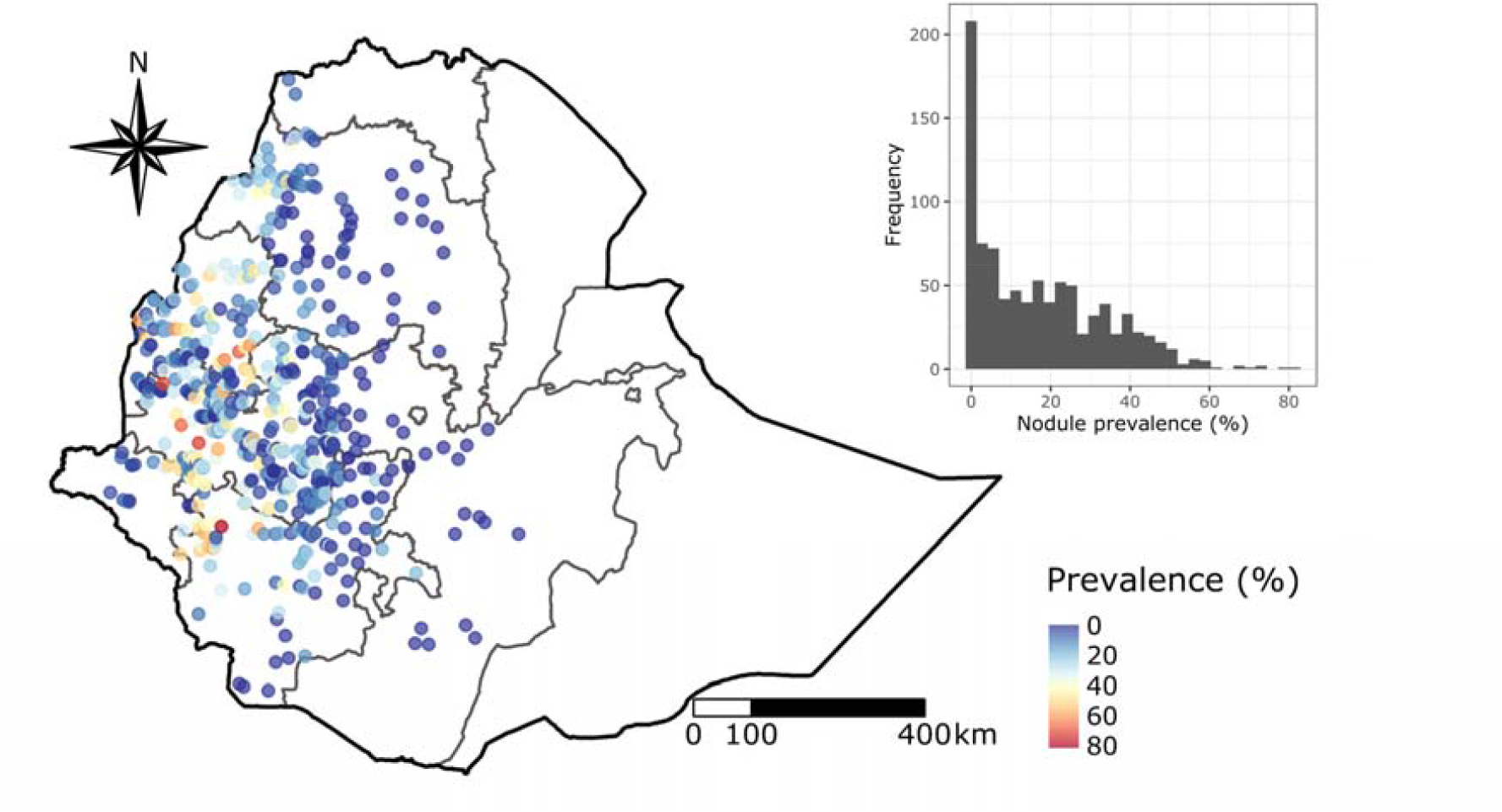
Sites and the nodule prevalence measured during Rapid Epidemiological Mapping of Onchocerciasis (REMO) in Ethiopia. The grey boundary on the map represents administrative regions. The inset figure shows the histogram of the prevalence.

### Environmental, climate, and socio-demographic variables

Variables relevant to *O. volvulus* infection prevalence and *Simulium* ecology based on published literature were assembled from different sources and were exported as a raster layer at a resolution of 1 km using Google Earth Engine (6, 36-39) (S1 Table). Raster layers with higher resolution were downsampled using a mean aggregation method, whereas the raster layers with lower resolution were resampled to align with 1 km resolution (40) to prepare a raster stack of uniform resolution. Raster data were processed using the *raster* package in R version 4.1.0 (41-43). Downloaded raster variables were reprojected to a standard projection, World Geodetic System 1984 (WGS84). The raster covariates were cropped to the boundary of Ethiopia (S1 Fig), and a raster stack of covariates was prepared. The measurement of different covariates at each sample site was extracted from the raster stack.

### Variable selection

Thirty-two variables were grouped into six major categories: elevation, temperature, precipitation, socio-demographic, hydrological, and vegetation (S1 Table), and the initial selection of covariates was conducted separately for each category. During the initial rounds of variable selection, multi-collinearity was assessed among the variables by calculating the Spearman’s rank correlation matrix and a variable inflation factor (VIF) for the linear model, including the variables, using the *GGally* and *car* packages in R (44, 45). Next, any variables with an absolute correlation coefficient less than 0.8 with other variables within the group were selected (46). For the set of covariates with a correlation coefficient greater than 0.8 and a VIF greater than 10, only one of the covariates was selected (33, 47). The VIF measures how easily a given predictor can be predicted from a linear regression based on other predictors. The predictor with the lowest VIF score was selected among the set of correlated covariates. The final covariates yielded a correlation matrix of less than 0.8 (S2 Fig) and a VIF factor of less than 10 (S2 Table). Based on this initial round of analysis, a set of 16 covariates were selected.

Model fit was assessed based on the Deviance Information Criterion (DIC) and Widely Applicable Information Criterion (WAIC) scores (48). We ran a univariate regression model and calculated the DIC and WAIC scores for the respective univariate models. A covariate yielding the least DIC and WAIC scores from each category was selected. Combinations of other variables were further explored if their inclusion further optimized the model fit scores. Eight covariates from the pool of 16 possible were selected for downstream geostatistical analysis (S3 Table).

### Geo-statistical modelling framework

A Bayesian geostatistical model was implemented using the Integrated Nested Laplace Approximation (INLA) approach, which has been reported to be computationally efficient for posterior distribution calculation and has been employed in recent large-scale geostatistical models (33, 46, 49, 50). Geostatistical approaches assume a positive spatial correlation between observations; i.e., the observations nearer to each other are more related than the farther ones. Information from neighboring pixels can then be utilized to allow smoothing of extreme values due to small sample sizes and give reliable and robust estimates from sparse data (33, 51). Further, the hierarchical structure of the model permits the estimation of covariate effects, spatial covariance structure, and the prediction of missing data (33). These models incorporate both fixed and random effects. The fixed effects determine the influence of covariates on *O. volvulus* infection prevalence, while the random effects account for the spatial variation that determines anomalous regions of high and low prevalence (46). This model can thus identify the relationship between infection prevalence data and several predictors and quantify spatial dependence via the covariance matrix of a Gaussian process facilitated by adding random effects to the observed locations (49).

### Model fitting

Conditional on the true prevalence *P*(*x*_*i*_) at location *x*_*i*_ = 1,2,3… .*n*, the number of cases (*Y*_*i*_) observed out of the total number of people tested (*N*_*i*_) were assumed to follow a binomial distribution.

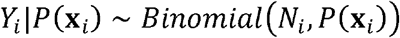

The log odds of prevalence is modeled as

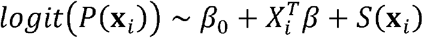

where *β*_0_ is the intercept, 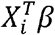 are the vectors of covariates and their coefficients. *S*(*x*_*i*_) is a spatial random effect with zero-mean Gaussian process following the Matérn covariance function which is defined by the equation:

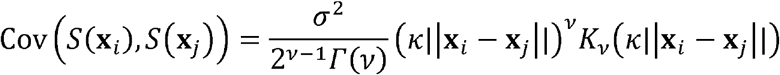

Here, *K*_*v*_ is the modified Bessel function of the second kind and order *v* > 0, *v* is the smoothness parameter, and *σ*^2^ is the marginal variance (46). *k* > 0 is the scaling parameter related to the practical range *ρ*, the distance at which the correlation between two points is approximately zero. However, if *ρ* = 8*vk*, at this range, the spatial correlation is close to 0.1 (9, 46). Default priors were used for the intercept parameter, effect parameters for the covariates, and the hyperparameters in the model as defined in Moraga, p. 35–37 (2).

### Accounting for excess zero prevalence

The binomial distribution is governed by only a single parameter which does not address overdispersion. To account for the excess zero prevalence in the data (Fig 1), zero-inflated binomial models (ZIB) Type 0 and Type 1 were also considered. There are structural zeros (prevalence reported to be zero based on reality) and sample zeros (prevalence reported to be zero based on chance) in any probability distribution (48, 52). Type 0 model considers only the structural zeros, while Type 1 considers both the structural and sample zeros. With ZIB Type 0 model, the probability density function for the observed cases is

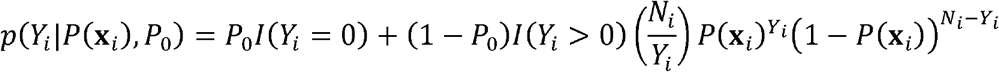

Here, *P*_0_ is the proportion of structural zeros, (1− *P*_0_) is the proportion of sample zeros, and *I*(*Y*_*i*_ = 0) is the indicator variable. When both structural zeros and sample zeros are considered, i.e., Type 1, the observations follow the probability density function:

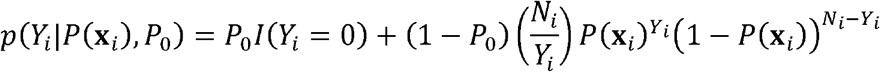

To determine the best fit model for the nodule prevalence data, model fit statistics (DIC and WAIC) were calculated for each model, viz. binomial, ZIB Type I, and ZIB Type 0.

### Mesh construction

We assume an underlying spatially continuous variable for the observed geostatistical data, which can be modeled with Gaussian random fields. We used the Stochastic Partial Differential Equation (SPDE) approach in the *INLA* package to fit a spatial model and to predict each variable of interest at an unsampled location (2, 53). An approximate solution to SPDE can be found using the finite element method. The finite element representation of the Matérn field is used as a linear combination of basis functions defined on a triangulation of domain D (54). Domain D is subdivided into a triangulated mesh which is formed first by placing the triangle’s vertices at the sample locations and then adding other vertices around the regions of spatial prediction.

We constructed the finite element mesh for SPDE approximation to the Gaussian process regression using the boundary of Ethiopia. Triangulation meshes with different cut-off parameters and the maximum length for the triangle inside and outside the boundary were tested for their model fit and computation cost. The mesh that yielded the lowest DIC and WAIC scores without significantly increasing computational cost was chosen.

### Cross-validation

K-fold cross-validation (with k = 10) was run to observe the differences in the predictive accuracy of the candidate models. Different measures of predictive accuracy were calculated by assessing the relationship between the predicted and observed prevalence in the validation dataset. In 10-fold cross-validation, the dataset is divided into ten random sections (or folds) (39, 46). Then, model validation runs are performed on each fold (10% of the data labeled the validation dataset) after fitting the data on the remaining nine folds (90% of the data labeled the training dataset). Thus, ten validation runs are performed for 10-fold cross-validation. During each validation run, both Spearman’s rank correlation coefficient and the Root Mean Square Error (RMSE) between the observed data and the predicted data for validation samples were calculated to assess accuracy.

### Prediction

The posterior distribution of prevalence was estimated at 5 km resolution, accounting for the effect of the variables and the spatial covariance structure. The covariate raster stack was aggregated to 5 km spatial resolution by taking either the mean or sum of the raster cells. The mean of raster cells was calculated for all continuous covariates except population count, for which the sum was calculated. Aggregated data were used to ease the computational burden associated with geospatial prediction at higher resolution. In addition, we calculated the aggregated mean, and the range of predicted prevalence values within a district/implementation unit (IUs). The predicted prevalence map was also used to assess the relationship with the environmental variables fitting the Generalized Additive Model (GAM) curve.

## Results

We formulated a Bayesian geostatistical model using INLA to estimate the nationwide pre-intervention prevalence of *O. volvulus* infection in Ethiopia. Nodule prevalence data from 916 unique geopositioned sites were combined with eight different environmental and socio-demographic covariates to construct the geostatistical model. Most of the prevalence data were from western Ethiopia, as eastern Ethiopia is largely unmapped for *O. volvulus* infection prevalence. The mean and the standard deviation of the observed prevalence across the sampling locations in Ethiopia was 17.24±16.32% ranging from 0 to 81.48%. There were 204 sites with zero prevalence (Fig 1).

### Model selection and fitting

Four different types of model were tested for the nodule prevalence data viz. binomial without spatial structure, binomial with spatial structure, ZIB type 1 and ZIB type 2, both with spatial structure. These were done without including any environmental and socio-demographic variables in the model. The binomial model that did not account for spatial effects showed higher DIC (9806.988) and WAIC (9816.581) scores (S3 Fig). The addition of spatial effect and accounting for zero inflation with a Type I zero-inflated binomial model decreased the DIC and WAIC scores to 5661.098 and 5916.715, respectively. Thus, ZIB Type I with spatial structure was chosen for modeling the prevalence data. To optimize the SPDE mesh, six different triangulation meshes with different parameters were tested for their model fit and computation cost (S4 Fig, S4 Table). The mesh C yielded the best model fit scores (DIC = 4538.12; WAIC = 4652.22). However, the mesh E yielded a comparable model fit (DIC = 4572.74; WAIC = 4710.781) but was computationally more efficient (45.38 s vs. 1667.33 s) and therefore, mesh E was chosen for fitting the model.

We selected environmental and socio-demographic variables based on the model fit scores of the univariate model. Isothermality was selected from the group of temperature variables, precipitation seasonality from the group of precipitation, and similarly, population density, distance to the nearest river, slope, and Normalized Difference Vegetation Index (NDVI) were selected from the group of socio-demographic, hydrological, and vegetation groups of covariates, respectively. Other combinations were also explored and the inclusion of covariates like soil moisture and flow accumulation further reduced the DIC and WAIC scores (S3 Table).

K-fold cross-validation (k = 10) was done for three different models: one without environmental covariates, one with six covariates, and the other with an additional two covariates (flow accumulation and soil moisture), which revealed that model 3 was superior to model 0 and 1 (S5 Fig). For model 3, calculating the Spearman rank correlation coefficient between the observed prevalence and the predicted prevalence ranged from 0.48 to 0.70 with a median of 0.66. Similarly, the RMSE ranged from 11.09 to 15.1, with a median of 13.18. This suggested a good model fit and accuracy for predictions across the validation datasets.

### Model parameters

The regression coefficients were estimated for each covariate included in the model. Since INLA is a Bayesian technique, the regression coefficients are a probability distribution rather than point estimates. A negative coefficient estimate implies a negative association of the variable with the prevalence and vice versa. The significance of the estimates was determined as described in Moraga et al. (55). The association was deemed significant only if both the 95% BCI values were below 0 for negative association and above 0 for positive association.

Out of 8 covariates considered for the final model, four covariates were significantly associated (based on 95% BCI) with *O. volvulus* infection prevalence (Table 1). Soil moisture was positively associated (0.0216, 95% BCI: 0.014 – 0.03) with *O. volvulus* infection prevalence, whereas distance to the nearest river (-0.015, 95% BCI: -0.025 – -0.005), precipitation seasonality (-0.017, 95% BCI: -0.032 – -0.001), and flow accumulation (-0.042, 95% BCI: -0.07 – -0.019) were negatively associated with *O. volvulus* infection prevalence. The regression coefficient of significant variables was at least one to two orders of magnitude greater than the non-significant ones.

**Table 1.** Mean coefficient estimates and 95% Bayesian credible interval (BCI) for the environmental and socio-demographic variables in the model. Regression coefficients for a particular covariate represent the change in *logit(P)* for a unit change in that covariate given that all other variables are kept constant.

| Variables | Regression coefficients |  |
| --- | --- | --- |
|  | Mean | 95% BCI |
| Distance to the nearest river | -0.01508 | (-0.0252, -0.005) |
| Soil moisture | 0.02158 | (0.0135, 0.0297) |
| Flow accumulation | -0.04225 | (-0.0703, -0.0186) |
| Precipitation seasonality | -0.01645 | (-0.0324, -0.0005) |
| Vegetation index | 0.00168 | (-0.0072, 0.0105) |
| Slope | 0.00097 | (-0.0071, 0.009) |
| Population density | 0.00005 | (-0.0003, 0.0003) |
| Isothermality | -0.00185 | (-0.0287, 0.0249) |
| Intercept | -1.94027 | (-4.9233, 1.0416) |
| **Hyper-parameters** |  |  |
| Zero probability parameter | 0.33345 | (0.327, 0.342) |
| $\theta_1$ for spatial field | -1.33992 | (-1.417, -1.286) |
| $\theta_2$ for spatial field | -0.09636 | (-0.17, -0.046) |
| 95% BCI includes 0.025 quantiles and the 0.975 quantiles of the probability distribution of the coefficients |  |  |

Hyperparameters defining the SPDE mesh were used to calculate the spatial effect and project the spatial field (S6 Fig). The spatial effect indicates the intrinsic spatial variability in the prevalence estimates, helping us understand the data’s spatial structure (47). Further, the spatial field also represents the spatial effect that was not accounted for by the covariates included in the model (55). The mean spatial field is higher in western Ethiopia while it is lower in central Ethiopia and eastern Ethiopia, along with the high standard deviation of the spatial field in the eastern parts.

### Model prediction

The predicted prevalence map shows spatial heterogeneity in *O. volvulus* infection prevalence in Ethiopia (Fig 2). Predicted *O. volvulus* infection prevalence is concentrated in the western parts of Ethiopia, with three to four hotspots in southwest Ethiopia. There is a relatively low prevalence of infection in eastern Ethiopia and near to zero prevalence in central Ethiopia. The range of predicted mean prevalence was 0.39 to 55.27%. Similarly, the lower limit of predicted infection prevalence ranged from 0 to 47.28%, while the upper limit of the predicted prevalence ranged from 1.41 to 65.32%. The correlation between the observed and the predicted prevalence was 0.71 (S7 Fig). Due to the geostatistical smoothing effect, some observations with higher prevalence were underestimated and vice-versa.

**Fig 2.**
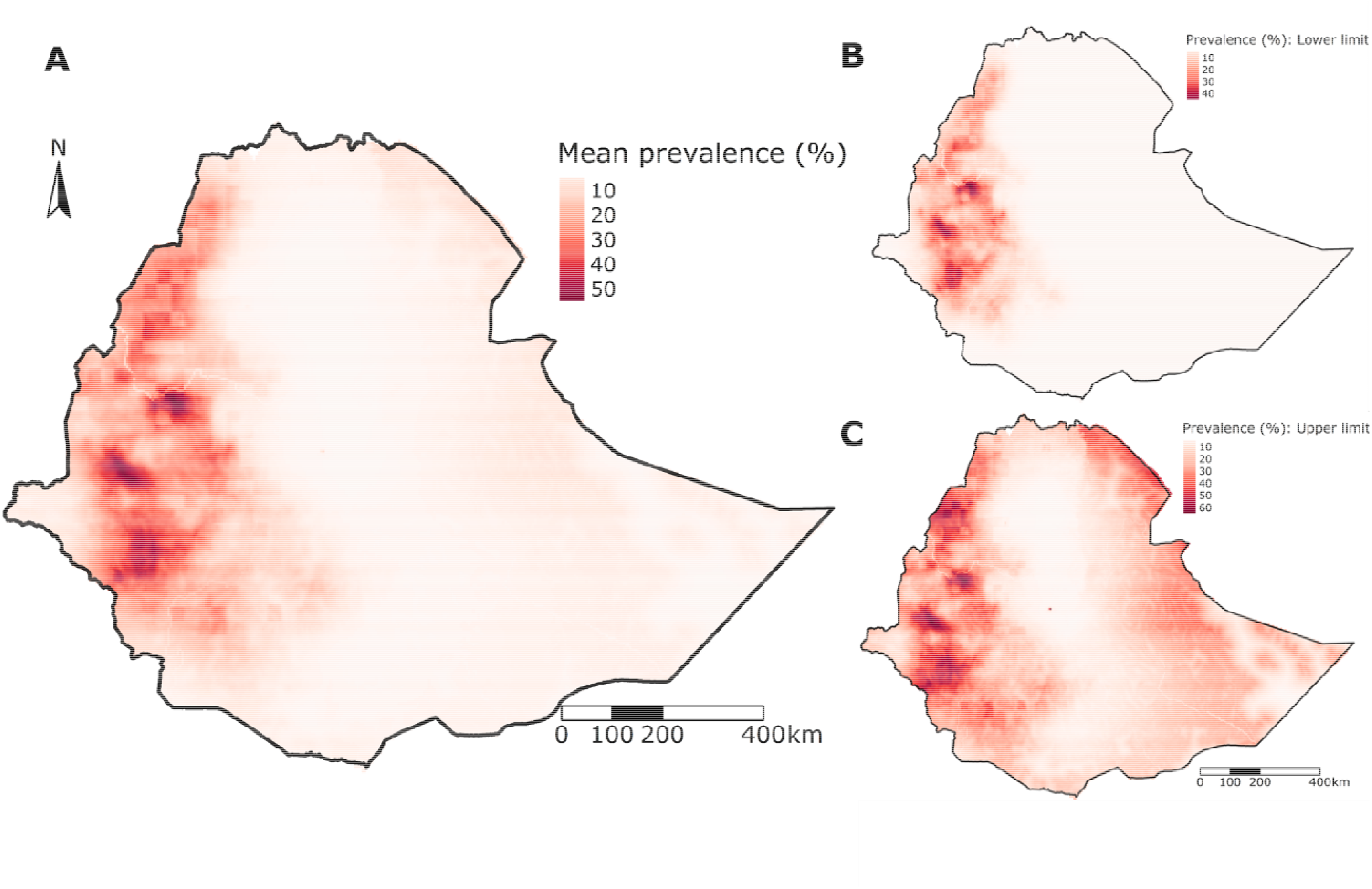
*Onchocerca volvulus* infection prevalence map in Ethiopia generated from the geostatistical model. The mean (A), the lower limit (B), and the upper limit (C) of *O. volvulus* infection prevalence. The prediction interval of the prevalence map is generated from the calculated 95% BCI of fitted prevalence values.

The uncertainty in the prevalence estimates was derived using the standard deviation of the posterior distribution. The uncertainty map shows that the presence of data influenced the uncertainty in the prevalence estimates; i.e., areas with the ground truth data have lower uncertainty (Fig 3A). The uncertainty was higher in eastern Ethiopia due to the lack of ground truth data from those sites. Most of central Ethiopia and some areas in eastern Ethiopia, regardless of the absence of the data, showed low prevalence with lower uncertainty (Fig 3B).

**Fig 3.**
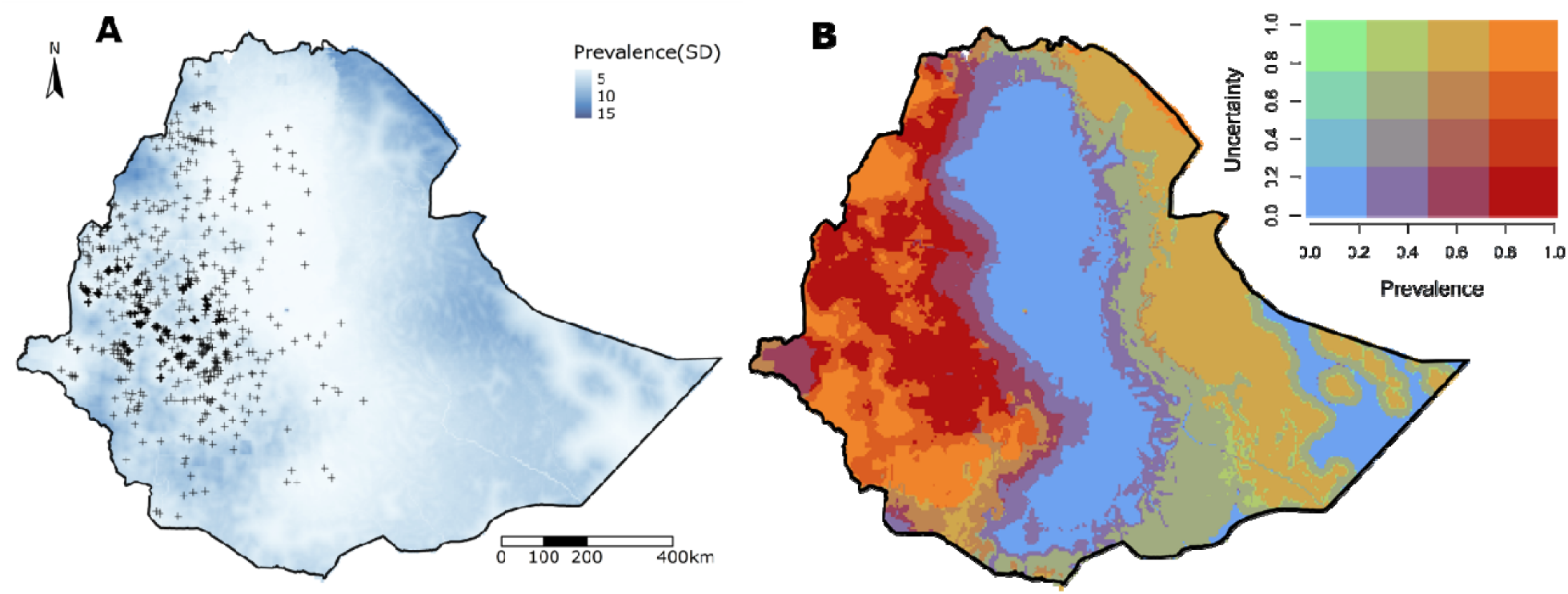
Uncertainty in the estimates of *O. volvulus* infection prevalence from the model. The standard deviation of the posterior distribution of prevalence (A) and the location of the observation are indicated by ‘+’ on the map. The bivariate map (B) shows both prevalence and the uncertainty estimates rescaled from 0 to 1.

There were areas with a high prevalence that had different levels of uncertainty in western Ethiopia. The regions with higher uncertainty almost always corresponded with sparse data from those regions.

A district-level map was created by aggregating the mean prevalence from pixels within the respective districts which represent implementation units (IUs) for MDAi (Fig 4). The aggregated mean prevalence for the first dozen of the most endemic districts were greater than 40%. The difference between the highest and the lowest estimated prevalence pixels (range of mean prevalence) within the districts was as high as 50.72% for a district within the Kemashi zone of Ethiopia.

**Fig 4.**
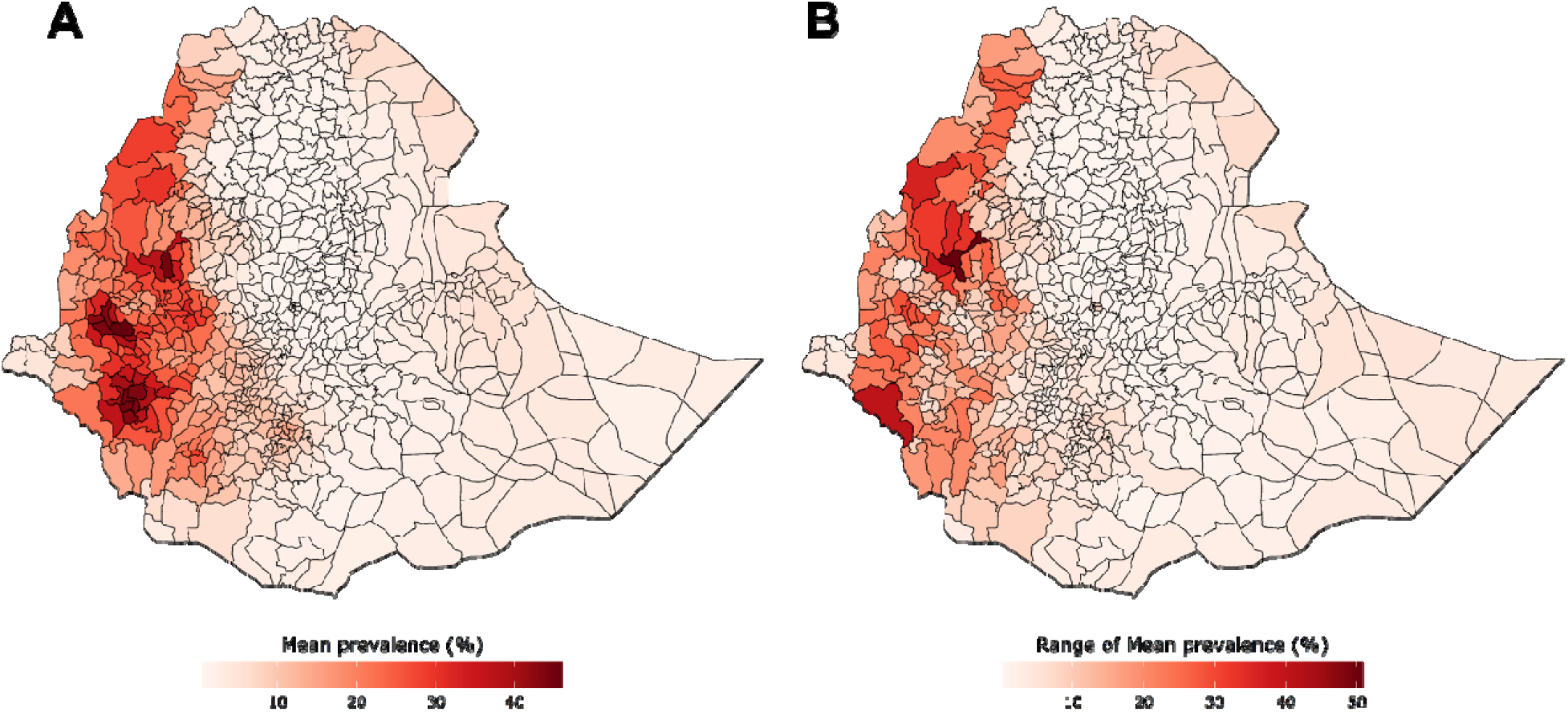
The aggregated mean prevalence and the range of the estimated mean prevalence within Ethiopian districts. The mean of the estimated prevalence of all the pixels within the district level border (**A**) and the range of the estimated prevalence within the district, i.e. the difference between the highest prevalence pixel and the lowest prevalence pixel (**B**), is shown.

### Relationship of environmental and socio-demographic covariates on the prevalence

A GAM curve was fitted between the predicted prevalence and the covariates used in the model to assess the relationship between them. The relationship profile of the predicted *O. volvulu* infection prevalence across the range of values of different covariates indicates which ecological conditions are suitable for onchocerciasis transmission (Fig 5). The relationship curve for the distance to the nearest river and the predicted prevalence shows the sharp decline in the *O. volvulus* infection prevalence to around 20-25 km, and the curve continues in the low prevalence region with increased uncertainty as distance increases from the nearest river (Fig 5). There wa almost a linear increase in the predicted prevalence with an increase in soil moisture up to around 18 mm.

**Fig 5.**
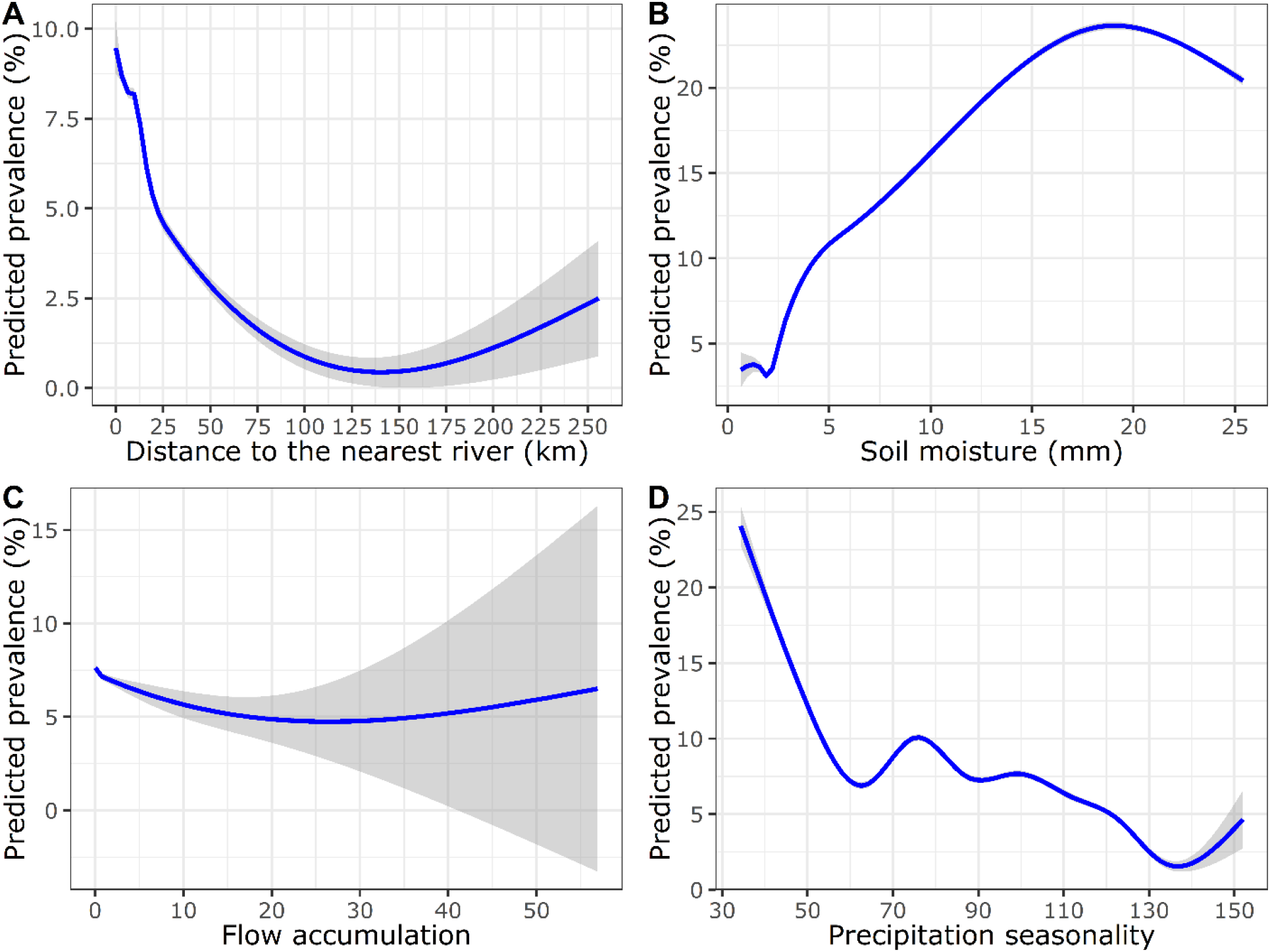
The relationship between the predicted mean prevalence with the significant environmental covariates in the regression model. The curve was fitted using a generalized additive model (GAM) using the smoothing function available in the *ggplot2* package. The shaded region around the curve represents the 95% confidence interval. Flow accumulation had a range of high magnitude compared to other covariates (values ranged from 0 to 100418). Thus, this variable was rescaled from 0 to 100 to make its range comparable with other variables.

There was a negative association with the flow accumulation with a considerable increase in uncertainty in the areas with high flow accumulation, i.e., larger rivers. Nevertheless, the areas with lower flow accumulation had a higher predicted prevalence than those with higher flow accumulation, suggesting the importance of intermediate-sized rivers to onchocerciasis epidemiology in Ethiopia. In addition, the relationship curve for the slope shows that a certain degree of slope is favorable for *O. volvulus* infection prevalence (S8 Fig). There is a similar response profile for population density where intermediate population density is favorable for onchocerciasis transmission. There was a steep decline in predicted prevalence with the initial increase in precipitation seasonality. However, there was a mixed non-linear response in the regions with precipitation seasonality from 60 to 130 mm.

## Discussion

We generated a country-level geospatial map of *O. volvulus* infection prevalence before the start of MDAi in Ethiopia, accounting for environmental and socio-demographic factors. The prevalence has been extrapolated to the country-level border of Ethiopia, including the eastern regions which were not mapped previously. Predicted prevalence in areas where people do not currently inhabit can indicate the risk of transmission should infected people establish communities. Prevalence was estimated using the pre-intervention nodule prevalence data and therefore represents the infection status before MDAi in Ethiopia. Thus, these predictions can act as a pre-control baseline map to on which decisions concerning new pre-MDAi mapping of likely hypoendemic areas that are not yet under MDAi can be prioritized and the effects of past interventions or of ecological changes at different locations can be assessed.

The predicted infection prevalence was found to be relatively low in the central parts of Ethiopia. This can be attributed to the presence of a significant geographical feature, the Great Rift valley. The elevated highlands along the center and lowland to the east of the Great Rift valley are characterized by low predicted prevalence. The land east of the valley is dry with few rivers (16, 20). On the other hand, the high elevation and slopes in western Ethiopia experience much higher rainfall, resulting in fast-flowing rivers, a specific requirement for blackfly breeding and development. The response profile for slope indicates that there is an optimal slope for the prevalence of *O. volvulus* infection that may be related to the flow characteristics optimal for blackfly breeding.

The spatial pattern of *O. volvulus* infection prevalence predicted across Ethiopia by this geospatial model was consistent with previously published prevalence maps that were based on REMO and other data (17, 19). It is noteworthy that in those earlier maps and in this geospatially predicted map there was a high level of spatial heterogeneity in infection prevalence, including heterogeneity within health districts (which are the implementation units for MDAi in Ethiopia). The difference between the highest and lowest prevalence pixel (range) within the districts was as high as 50%. A study in Cameroon reported that hypoendemic areas could sustain low-grade transmission and, therefore, might cause rapid recrudescence in neighboring meso- and hyperendemic areas where the transmission has been successfully controlled (56). Given that much of the unmapped onchocerciasis endemic areas of Ethiopia is hypoendemic, these areas must be identified and treated for elimination of transmission to be reached. Hence, we need to consider the spatial heterogeneity within and between the intervention units while planning the elimination programs.

We used a bivariate map to visualize estimated prevalence and the associated uncertainty (Fig 4). The presence/absence of data influences this uncertainty map; i.e., areas with ground truth data have lower prediction errors. This is expected in geostatistical models as they depend on the Euclidean distances between the reported observations (39). Thus, the uncertainty map can indicate where additional data would reduce the overall prediction error of the prevalence map, particularly in the areas with higher prevalence. This can be used to identify regions that might benefit from targeted re-mapping or elimination mapping efforts (3). For example, there are areas with higher prevalence in the west but varying uncertainty. The areas with high uncertainty could be targeted for re-mapping. Similarly, there are areas in the east with both low prevalence and lower uncertainty, i.e., with higher confidence, and thus, do not need to be re-mapped.

### Ecological features associated with *O. volvulus* infection prevalence

The major environmental factors significantly associated with infection prevalence were distance to the nearest river, soil moisture, precipitation seasonality, and flow accumulation. As expected, there was a negative association between the distance to the nearest river and predicted prevalence. Onchocerciasis has long been recognized as being higher in communities near rivers and this correlation, which has been reported in prior geospatial modeling studies (19, 37, 39), is driven by blackfly breeding and development requirements for fast-flowing rivers, such that villages can be categorized epidemiologically as first, second, or third-line villages based on their proximity to vector breeding sites (39, 57, 58).

The relationship curve between the predicted prevalence and the distance to the nearest river shows that there is an initial rapid decline in prevalence followed by a less rapid decline as the distance from the river increases, and the curve asymptotes to a very low prevalence with increased uncertainty as the distance exceeds 100 km. A rapid decline in blackfly biting rate at increasing distance from a river breeding site, based on vector biting rate data collected in northern Cameroon over three years, has been reported previously (59). Similarly, a mark-recapture study found a logarithmic decline in the proportional fly biting density as the distance increased from the marking site (60), and a mark-release-recapture study in Ghana in West Africa reported the average flight range of *S. damnosum* may be as high as 27 km (58). While Ethiopia is host to several different competent blackfly vector species, the part of the curve where the change in slope declines is consistent with this estimated flight range viz., ∼20-25 km. However, the curve does not reach its lowest point until 100 km, suggesting that the parasites could be transmitted beyond the average dispersal range of an individual blackfly. This could be because the dispersal range for gravid blood-seeking and ovipositing female blackflies has been reported to be greater than the average dispersal range at around 60-100 km from the river (58). In addition, wind-assisted long-distance migration of blackflies of hundreds of km and transmission due to the human migration have also been reported (61-63). Thus, this study supports that longer range migration is likely and that it also likely contributes to *O. volvulus* transmission.

We observed a positive association between soil moisture and *O. volvulus* infection prevalence. Soil moisture is high in areas with high precipitation or near water bodies, including rivers—i.e., where there are suitable blackfly breeding sites. Soil moisture is an indicator of agricultural suitability, and agricultural areas have historically known to have high prevalence of *O. volvulus* infection (19, 37, 64). Agricultural lands and farms in these areas tend to be near rivers for easy irrigation. Therefore, the increased prevalence of *O. volvulus* infection among people involved in agriculture and farming (37, 65, 66) is presumably because these workers are generally outdoors, often in proximity to rivers, and thus experience increased exposure to blackflies (7, 9, 67).

Flow accumulation is used in hydrogeology as a proxy for river grades and represents the cumulative number of cells in a raster object that flow into a given cell: high flow accumulation represents large rivers, and lower flow accumulation represents secondary rivers and their tributaries. It has been used to map onchocerciasis hotspots in hypoendemic settings of the Democratic Republic of Congo (68). In this study, flow accumulation was negatively associated with *O. volvulus* infection prevalence, meaning that the infection was more common in the communities near the secondary rivers and tributaries than the large rivers. The primary vectors of onchocerciasis in Ethiopia are *S. damnosum s*.*l*. and *S. neavei* (69). In a study describing the ecological study of West African *Simulium* spp., *S. damnosum s*.*l*. were found in rivers of medium width with a lower flow accumulation than the large size rivers (6). Furthermore, the important characteristic of *S. neavei* is the obligatory phoretic association of larvae with the freshwater crabs which are more common in sheltered smaller river streams in the forests than in larger rivers (70, 71). The forested riverine areas, where *S. neavei* are found, is a dominant ecotype in southwestern midland and highlands of Ethiopia, where onchocerciasis is highly endemic and from where most of the data were collected for this study (9, 72).

Similarly, precipitation seasonality was also negatively associated with the predicted prevalence, i.e., the prevalence was high in the areas with lower precipitation seasonality. Areas with high precipitation seasonality might have ephemeral rather than perennial streams. If the breeding sites are ephemeral, blood-feeding by blackflies would only happen during some part of the year, lowering the annual biting rate, a key parameter in *O. volvulus* transmission (73-75). Southeast Ethiopia, characterized by low prevalence of *O. volvulus* infection, has high seasonality in precipitation that is characterized by two short wet seasons with a dry period in between (76). However, the southwestern areas where the disease is most endemic have low precipitation seasonality and high annual precipitation (77).

As one would expect, the environmental factors that were significantly associated with *O. volvulus* infection prevalence are all exert strong influence on vector breeding and thus impact blackfly density and biting rates. The implication of this strong association between determinants of vector breeding and infection prevalence implies that spatial variation in vector breeding drives the spatial variation of *O. volvulus* infection prevalence in Ethiopia and that the geospatial model we present here, based on nodule prevalence data, is also predictive of vector distribution. This suggests that ongoing climate change, which is affecting the pattern of precipitation (78, 79), and other anthropogenic environmental changes such as changing river flow with the construction of river dams for hydroelectricity or irrigation might significantly change vector distribution and thus the spatial occurrence of the disease (80, 81). The impacts of these changes could be modeled using this approach.

### Model limitations and recommendations

The geospatial model we report here incorporates different environmental and socio-demographic variables that are known to influence the transmission and prevalence of *O. volvulus* infection and the distribution of blackflies. However, the data incorporated in the model do not include all factors that may be epidemiologically relevant, such as direct/indirect interventions affecting infection prevalence and human behaviors that may increase or decrease the risk of infection. The non-uniform mean spatial field across the triangulation mesh shows that there might be some effects that are unaccounted for by the model (S6 Fig), and the possibility remains that an unidentified covariate that closely resembles the spatial field might aid in explaining the spatial variation in prevalence. In addition, the inclusion of blackfly distribution maps based on the identification of breeding sites and their productivity might improve the model fit. Unfortunately, such data are not available for Ethiopia.

Some variables that we expected may correlate with prevalence, such as human host population density and vegetation, were not shown to be significantly associated in these analyses. Blackflies are not usually reported to be found in dense urban environments and, similarly, vegetation cover is essential for blackfly breeding and thus *O. volvulus* transmission (37, 64, 82). We suggest that the lack of association might be because the country-wide spatial scale neutralizes factors that impact prevalence at a smaller geographic scale. Therefore, targeted spatial analysis in regions with differences in vegetation near rivers or with differences in rural-urban indices (83) might be helpful to explore the effects of these variables on infection prevalence. Furthermore, *O. volvulus* transmission is highly dynamic, not just spatially but also temporally. Extending the current spatial model to a spatio-temporal model might improve the model fit, which requires both the prevalence and the covariate data at sufficient temporal resolution.

We could not include prevalence measures based on other diagnostic methods for Ethiopia because fine-scaled prevalence data based on mf counts from skin snips or antibody tests (Ov16) were not available. However, these data could be used as an alternative to, or in addition to, nodule prevalence. Combining data across methods is challenging, as correlations between mf counts and nodule counts can be highly variable (however, see (18)) and the correlation between these measures and Ov16 seropositivity prevalence is unclear. Nevertheless, the map presented here could be used by onchocerciasis elimination programs to direct resources for elimination mapping because elimination mapping of any disease can be expensive (3), and the method described here may be an inexpensive first step that can extrapolate country-wide prevalence from existing data and thus better target re-mapping efforts.

## Conclusion

Onchocerciasis programs have transitioned from control of onchocerciasis as a public health problem to elimination of *O. volvulus* transmission, triggering the need to develop new tools to more efficiently prioritize decisions concerning elimination mapping and interventions in hypoendemic foci that were not previously targeted for intervention. To this end, we have generated a baseline pre-intervention prevalence map for the whole of Ethiopia using geospatial modelling that is based on pre-intervention nodule prevalence data and spatial variation in different environmental and socio-demographic factors. We extrapolated existing historical nodule prevalence measures to previously unmapped regions of Ethiopia and quantified uncertainty in predicted prevalence. This map could be used as an aid to decision making on where and how to (a) extend elimination mapping into areas identified as likely hypoendemic foci and (b) prioritize the allocation of scarce health system resources to areas most likely to benefit from that allocation. Furthermore, this study found that hydrological variables such as distance to the nearest river, soil moisture, precipitation seasonality, and flow accumulation were significant in describing the spatial heterogeneity of *O. volvulus* infection in Ethiopia. All these ecological features are related to the suitability of an area for vector breeding, movement, biting behavior, and density, leading to the conclusion that vector suitability and movement are the primary determinants of the spatial distribution of *O. volvulus* infection in Ethiopia. Consequently, changes in these ecological features due to anthropomorphic changes in climate, agriculture, vegetation type (e.g., slash-and-clear), or construction of hydroelectric or irrigation dams might significantly alter the occurrence of the disease. We suggest, therefore, that the importance of these vector-related ecological factors in determining infection distribution and intensity reaffirms that inclusion of vector control could augment current interventions based primarily on prophylactic chemotherapy.

## Data Availability

The prevalence data underlying the results presented in the study are available from the ESPEN website (https://espen.afro.who.int/tools-resources/download-data). Details on the sources for the environmental variables used are in the Supplementary Information.

## Acknowledgements

The authors wish to acknowledge Mr Sindew M. Feleke at the Ethiopian Public Health Institute (EPHI), Dr Moses Katabarwa at the Carter Center for assistance in navigating the data sources and providing insights into Ethiopian control programs, and Dr Katie Crawford and Mr Haylo Roberts for helpful discussion during the analysis.

