## Supplemental information for "Geospatial modeling of pre-intervention prevalence of *Onchocerca volvulus* infection in Ethiopia as an aid to onchocerciasis elimination"

### Supplementary information

**S1 Table. Information on environmental and socio-demographic covariates considered for the geospatial analysis.**

| **S.N.** | | |  | **Categories** | | **Covariates** | | **Resolution** | | **Source** | | **Time** | | **Unit** | | **References** |
| --- | --- | --- | --- | --- | --- | --- | --- | --- | --- | --- | --- | --- | --- | --- | --- | --- |
| 1 |  | Temperature | | | BIO1 = Annual Mean Temperature | | 30 arc second (~1 km) | | WorldClim V1 Bioclim | | 1970-2000 | | °C | |  | |
|  |  |  | | | BIO2 = Mean Diurnal Range (Mean of monthly (max temp - min temp)) | |  | |  | |  | | °C | |  | |
|  |  |  | | | BIO3 = Isothermality (BIO2/BIO7) (×100) | |  | |  | |  | | % | |  | |
|  |  |  | | | BIO4 = Temperature Seasonality (standard deviation × 100) | |  | |  | |  | | °C | |  | |
|  |  |  | | | BIO5 = Max Temperature of Warmest Month | |  | |  | |  | | °C | | [1] | |
|  |  |  | | | BIO6 = Min Temperature of Coldest Month | |  | |  | |  | | °C | |  | |
|  |  |  | | | BIO7 = Temperature Annual Range (BIO5-BIO6) | |  | |  | |  | | °C | |  | |
|  |  |  | | | BIO8 = Mean Temperature of Wettest Quarter | |  | |  | |  | | °C | |  | |
|  |  |  | | | BIO9 = Mean Temperature of Driest Quarter | |  | |  | |  | | °C | |  | |
|  |  |  | | | BIO10 = Mean Temperature of Warmest Quarter | |  | |  | |  | | °C | |  | |
|  |  |  | | | BIO11 = Mean Temperature of Coldest Quarter | |  | |  | |  | | °C | |  | |
| 2 |  | Precipitation | | | BIO12 = Annual Precipitation | | 30 arc second (~1 km) | | WorldClim V1 Bioclim | | 1970-2000 | | mm | |  | |
|  |  |  | | | BIO13 = Precipitation of Wettest Month | |  | |  | |  | | mm | |  | |
|  |  |  | | | BIO14 = Precipitation of Driest Month | |  | |  | |  | | mm | |  | |
|  |  |  | | | BIO15 = Precipitation Seasonality (Coefficient of Variation) | |  | |  | |  | | coefficient of variation | | [1] | |
|  |  |  | | | BIO16 = Precipitation of Wettest Quarter | |  | |  | |  | | mm | |  | |
|  |  |  | | | BIO17 = Precipitation of Driest Quarter | |  | |  | |  | | mm | |  | |
|  |  |  | | | BIO18 = Precipitation of Warmest Quarter | |  | |  | |  | | mm | |  | |
|  |  |  | | | BIO19 = Precipitation of Coldest Quarter | |  | |  | |  | | mm | |  | |
| 3 |  | Elevation | | | Digital Elevation Model (DEM) | | 90 m | | CGIAR-SRTM | | 2000 | | meters | | [2] | |
|  |  |  | | | Slope | | 3 arc second (~100m) | | WorldPop SRTM | | 2000 | | topographic slope in degree | |  |  |
| 4 |  | Vegetation indices | | | NDVI*^+^ | | 1 km | | MOD13A2 V6, NOAA | | 2003-2011 | | NA | | [3] | |
|  |  |  | | | EVI* | | 5 km | | Malaria Atlas Project | | 2003-2011 | | NA | | [4] | |
| 5 |  | Hydrological data | | | Flow accumulation^+^ | | 30 arc sec (~1 km) | | HydroSHEDS, WWF | | 2000 | | Number of cells | | [5, 6] | |
|  |  |  | | | Drainage direction | |  | |  | | 2000 | | N.A. | |  |  |
|  |  |  | | | Distance to the nearest river (derived from water lines in DIVA-GIS) | | 1km | |  | | 2003 | | km | | [7] | |
|  |  |  | | | Soil moisture* | | 0.25 degrees (25 km) | | NASA-USDA Global soil moisture | | 2010-2012 | | mm | | [8] | |
| 6 |  | Socio-demographic | | | Population density* | | 30 arc second (~1 km) | | Gridded Population of World Version 4 (GPWv4) | | 2000-2012 | | density | | [9] | |
|  |  |  | | | Nighttime lights* | | 100 m | | WorldPop (Resampled DMSP, OLS) | | 2000-2011 | | 0-6300 | | [10] | |
|  |  |  | | | Nighttime lights* | | 100 m | | WorldPop (Resampled VIIRS) | | 2012-2016 | | nanoWatts/cm2/sr. | |  |  |
|  |  |  | | | Improved housing (prevalence)* | | 30 arc second (~1 km) | | Malaria Atlas Project | | 2000, 2015 | | % | | [11] | |
| 7 |  | Administrative boundaries | | | Administrative boundaries for Ethiopia and Africa | |  | | Database of Global Administrative Areas (GADM) | | 2012 | | NA | | [12] | |

*A static mean layer was derived for the variables which had raster layers at multiple time points

^+^ Flow accumulation and NDVI were rescaled from 0 to 100 due to their high range.

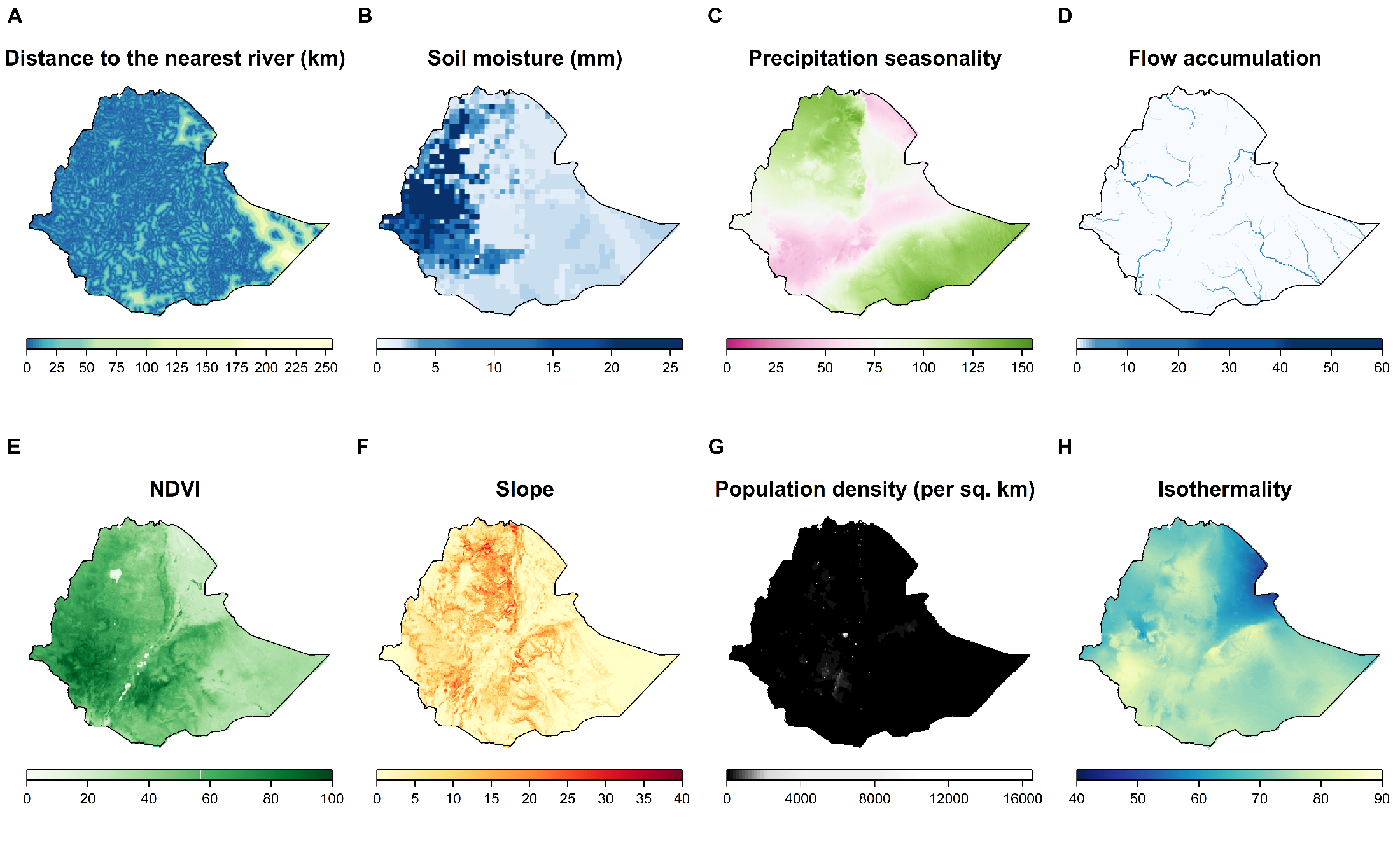

**S1 Fig. Socio-demographic and environmental covariates used in the geostatistical model.** The raster layers are masked to the border of Ethiopia. Flow accumulation and NDVI are rescaled from 0 to 100. NDVI: Normalized Difference Vegetation Index

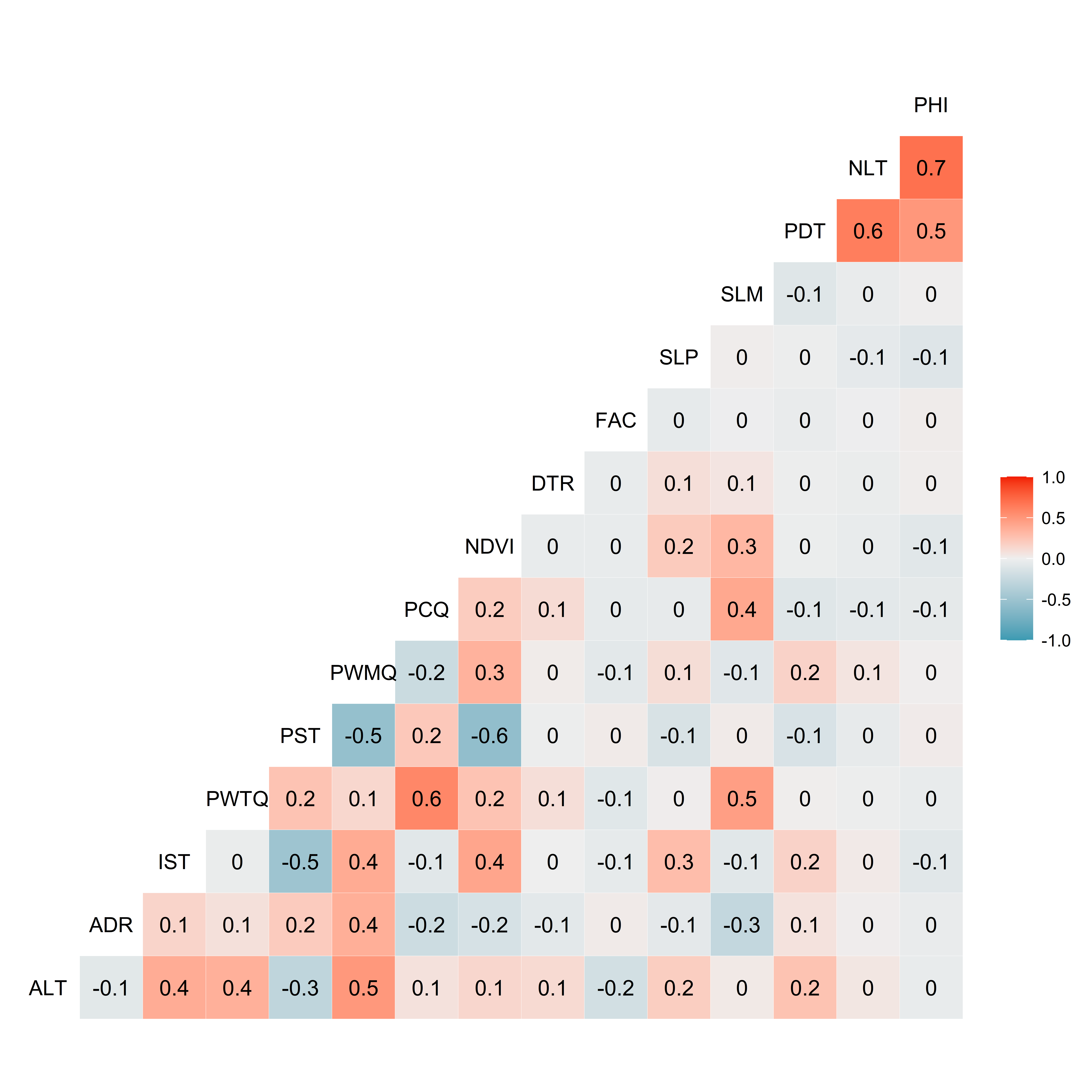

**S2 Fig. Correlation matrix of the 16 environmental and socio-demographic variables selected after the initial round of covariate selection.** Spearman's rank correlation coefficient was estimated assuming the non-normality of the data, and the correlation coefficient for each pair of covariates was below 0.8. ELV: elevation; ADR: annual diurnal range; IST: isothermality; PWTQ: precipitation wettest quarter; PST: precipitation seasonality; PWMQ: precipitation warmest quarter; PCQ: precipitation coldest quarter; NDVI: normalized difference vegetation indices; DTR: Distance to the nearest river; FAC: flow accumulation; SLP: slope; SLM: soil moisture; PDT: population density; NLT: night lights; PHI: prevalence of housing improvement

**S2 Table. Variable inflation factor (VIF) for 15 covariates selected during the initial round of variable selection.**

| **S.N.** | **Variables** | **Variable inflation factor (VIF)** |
| --- | --- | --- |
| 1 | Elevation | 2.603145 |
| 2 | Annual diurnal range | 2.391153 |
| 3 | Isothermality | 2.01164 |
| 4 | Precipitation wettest quarter | 3.125196 |
| 5 | Precipitation seasonality | 3.962958 |
| 6 | Precipitation warmest quarter | 3.385919 |
| 7 | Precipitation coldest quarter | 1.870921 |
| 8 | NDVI | 2.672129 |
| 9 | Distance to the nearest river | 1.057526 |
| 10 | Flow accumulation | 1.036067 |
| 11 | Slope | 1.163392 |
| 12 | Soil moisture | 1.5485 |
| 13 | Population density | 1.930984 |
| 14 | Night lights | 2.452299 |
| 15 | Prevalence of housing improvement | 2.059249 |

**S3 Table. Information criterion scores for each potential explanatory variable(s).** A univariate spatial model was fitted to each variable, and the DIC and WAIC scores were calculated. Variables were grouped into the category they represented, and the variable with the least DIC and WAIC scores were selected. Other variables were explored after combining the selected variables.

| **Model type** | **Covariates group** | **Covariates** | **WAIC** | **DIC** |
| --- | --- | --- | --- | --- |
| Univariate models | 1 | Elevation | 4870.679555 | 4705.188298 |
|  |  | Slope | 4815.251927 | 4661.838606 |
|  | 2 | Annual diurnal range | 4826.705701 | 4671.029979 |
|  |  | Isothermality | 4771.03046 | 4630.35894 |
|  | 3 | Precipitation wettest quarter | 4798.416596 | 4647.965794 |
|  |  | Precipitation warmest quarter | 4870.29998 | 4709.206525 |
|  |  | Precipitation seasonality | 4694.56486 | 4551.345871 |
|  |  | Precipitation coldest quarter | 4992.524611 | 4798.803025 |
|  | 4 | NDVI | 4713.170199 | 4577.706866 |
|  | 5 | Night lights | 4857.113257 | 4692.447611 |
|  |  | Improvement in housing | 4821.227795 | 4657.575238 |
|  |  | Population density | 4763.297884 | 4622.747077 |
|  | 6 | Flow accumulation | 4764.449976 | 4623.255219 |
|  |  | Distance to the nearest river | 4694.979031 | 4565.819538 |
|  |  | Soil moisture | 4753.415409 | 4616.727082 |
| Models with the combination of selected covariates | 7 | Slope + Isothermality + Precipitation seasonality + NDVI + Poppulation density + Distance to the nearest river | 4857.92 | 4688.2 |
|  | 8 | Slope + Isothermality + Precipitation seasonality + NDVI + Poppulation density + Distance to the nearest river + Flow accumulation + Soil moisture | 4749.86 | 4612.4 |

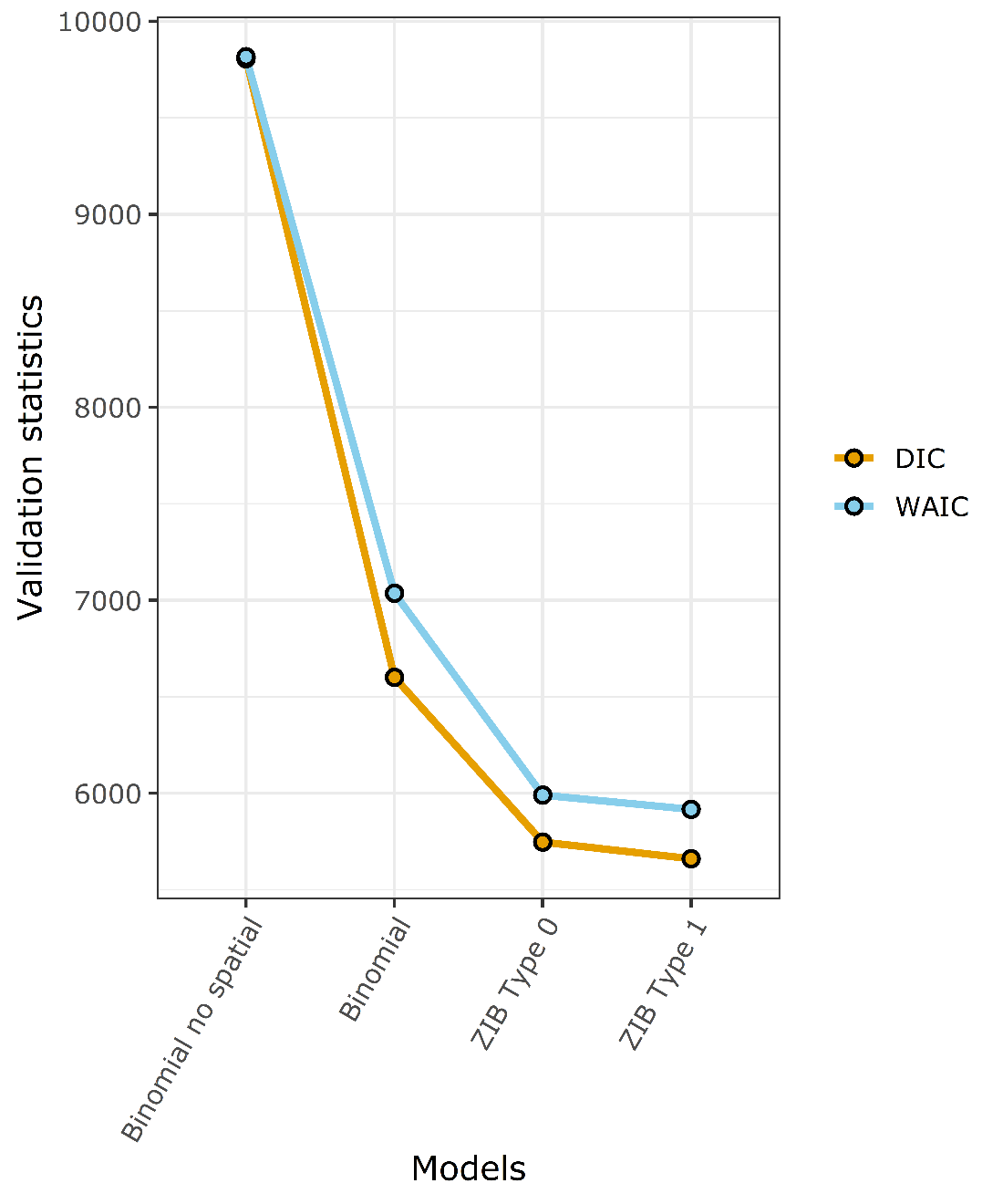

**S3 Fig. Changes in the model fit statistics for different types of models.** Type 1 zero-inflated binomial distribution yielded the lowest AIC and WAIC scores suggesting the best model fit.

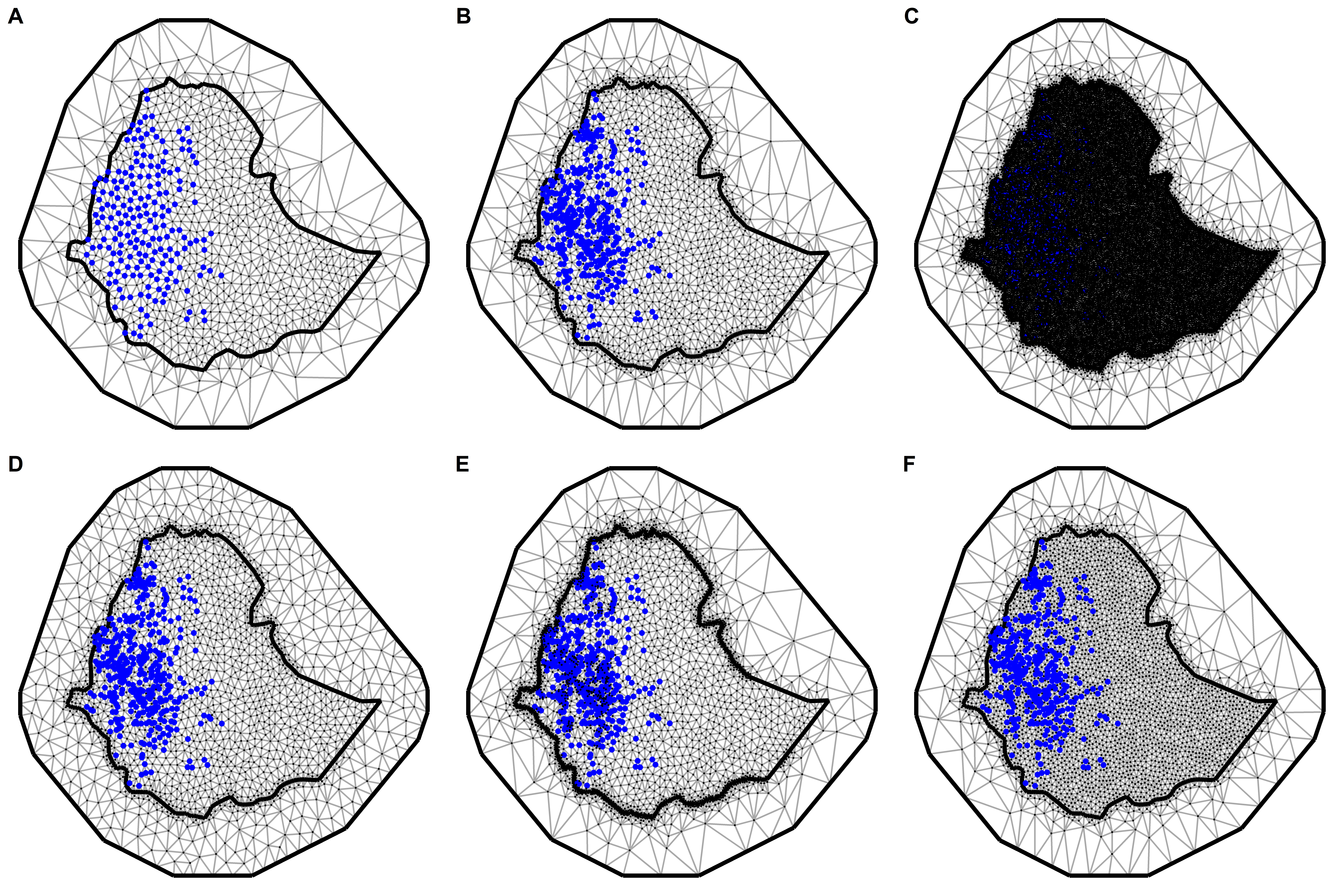

**S4 Fig. Different triangulated SPDE meshes considered for the analysis.** The country boundary for the prediction region and the location of the observations (blue points) are shown on the triangulation mesh. The finer mesh is within the country boundary, while the coarser mesh is present outside the country boundary in the buffer region.

**S4 Table. The parameters, model fit scores, and the computational cost for different meshes shown on S4 Fig.** A reasonable improvement in model fit was achieved with Mesh E without compromising the computational cost. Thus, Mesh E was chosen to represent the Matérn field. The time taken for the computation is based on a machine with Intel i7, 3.8 GHz processor.

|  | **Mesh parameters** | | |  |  | **Model fit scores** | |
| --- | --- | --- | --- | --- | --- | --- | --- |
| **Mesh** | **Cut-off** | **Inner triangle maximum length** | **Outer triangle maximum length** | **Number of vertices** | **Time taken (s)** | **DIC** | **WAIC** |
| A | 0.3 | 0.5 | 5 | 878 | 16.69 | 5573.18 | 5834.05 |
| B | 0.1 | 0.5 | 5 | 1827 | 27.58 | 4818.08 | 5010.92 |
| C | 0.01 | 0.1 | 5 | 27882 | 1667.33 | 4538.12 | 4652.22 |
| D | 0.1 | 0.5 | 1 | 2004 | 30.84 | 4946.56 | 5150.15 |
| E | 0.03 | 0.5 | 5 | 3931 | 45.38289 | 4572.74 | 4710.781 |
| F | 0.1 | 0.3 | 5 | 3197 | 70.86 | 4892.32 | 5106.08 |

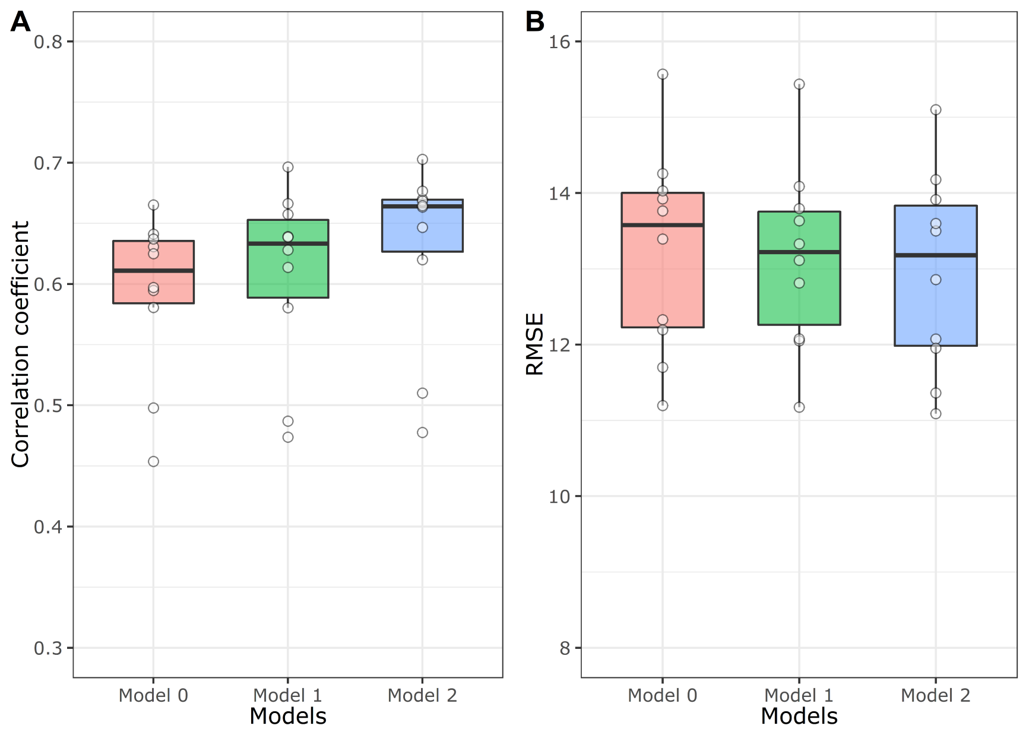

**S5 Fig. Boxplot showing cross-validation statistics obtained from 10-fold cross-validation from the three different geostatistical models.** Spearman's rank correlation coefficient (A) and RMSE (B) were calculated between the predicted and the observed prevalence of each validation set during each cross-validation run. Model 0 is the model with only intercept and the spatial field, model 1 consists of six variables (slope, isothermality, precipitation seasonality, NDVI, population density, and distance to the nearest river) with intercept and the spatial field, and model 2 consists of everything in model 1 with two additional variables (flow accumulation and soil moisture).

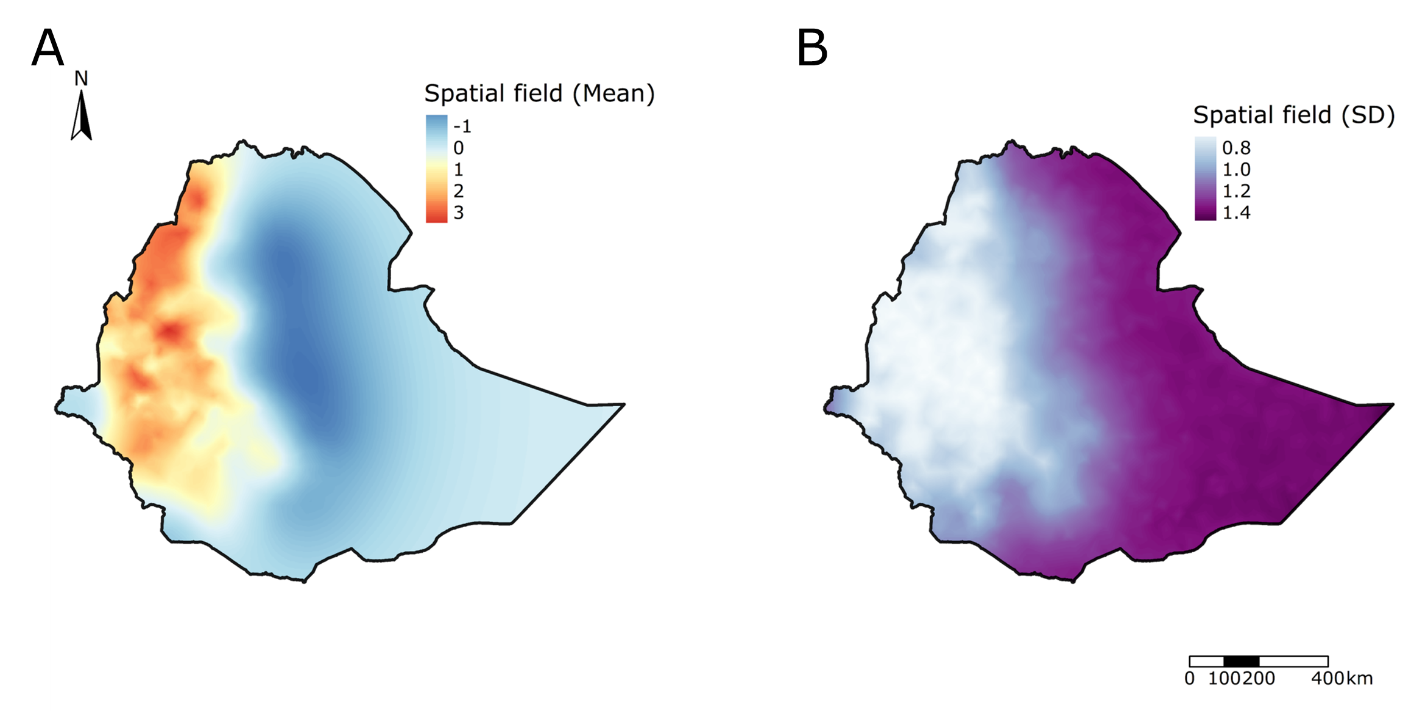

**S6 Fig. The spatial field's posterior mean (A) and standard deviation (B) from the stochastic partial differential equation (SPDE) mesh.** The spatial field is higher in western Ethiopia. While the spatial field is lower in eastern Ethiopia, the standard deviation of the spatial field is higher.

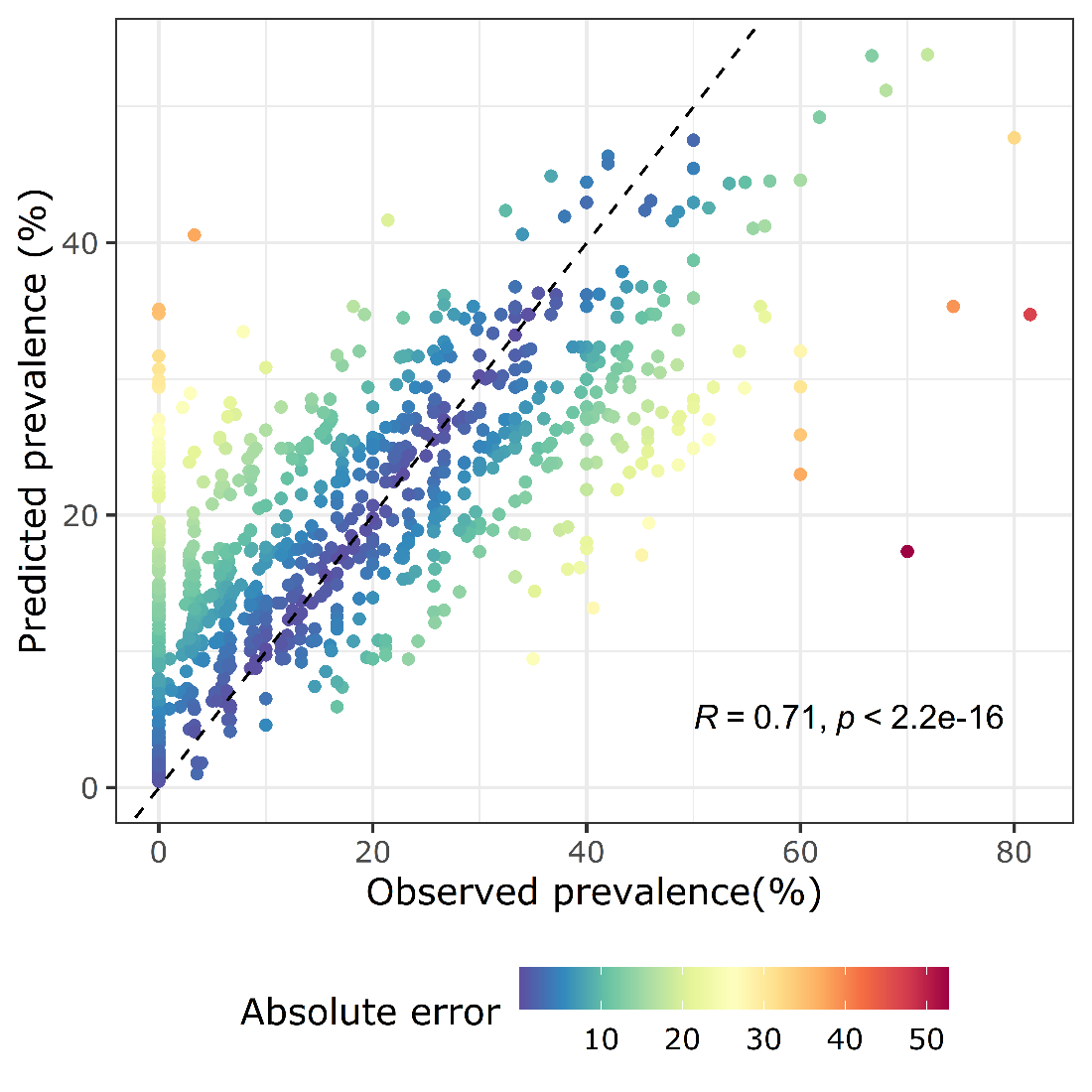

**S7 Fig. Correlation between the observed and predicted prevalence.** The dashed line is the expectation for perfect correlation. The Spearman's rank coefficient and associated p-value are shown on the bottom left of the plot. Points are colored by the absolute difference between the observed and the predicted prevalence.

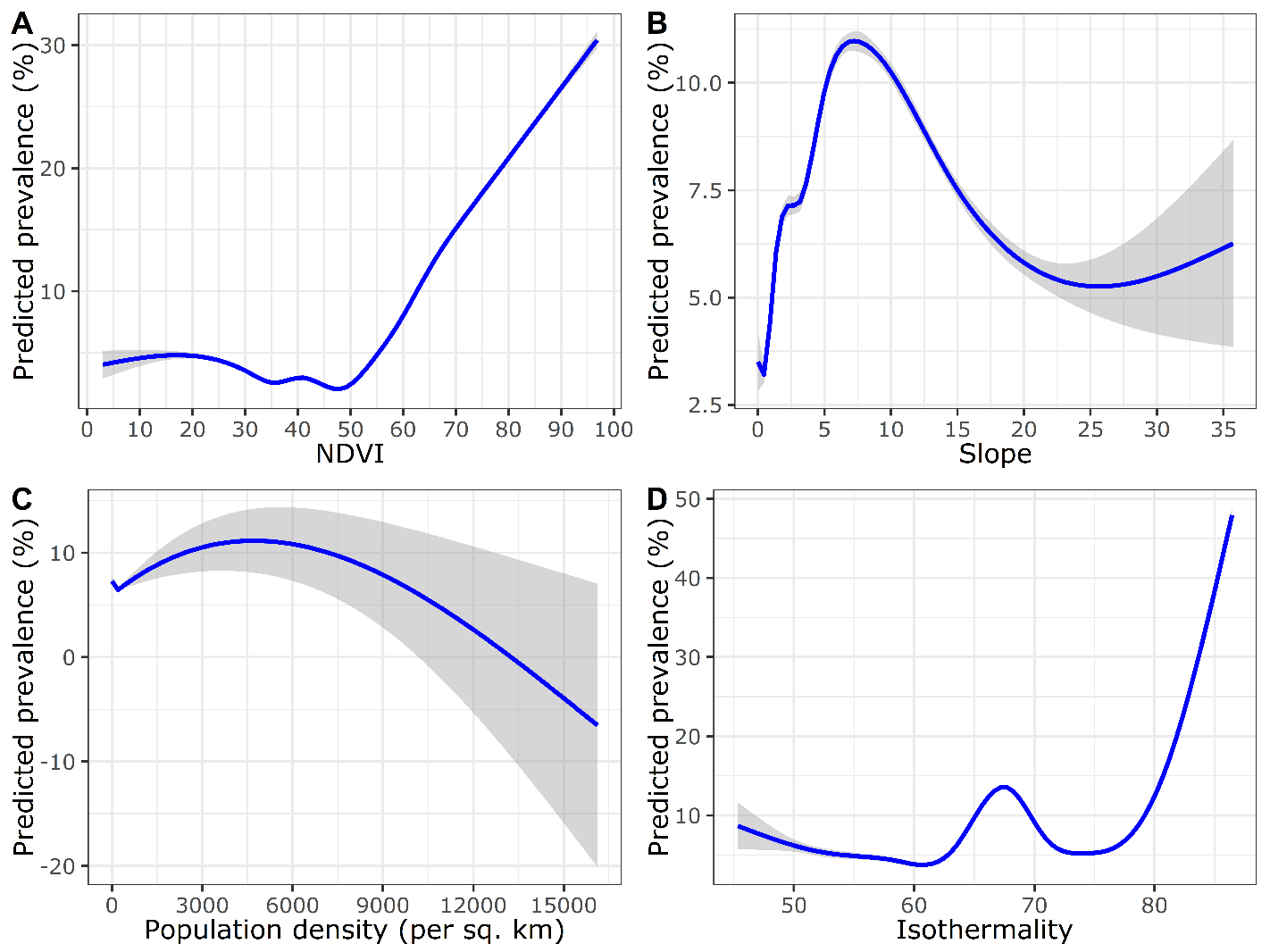

**S8 Fig. The relationship between the predicted mean prevalence with the non-significant environmental and socio-demographic covariates in the regression model.** The curve was fitted using a generalized additive model (GAM) using the smoothing function available in the *ggplot2* package. The shaded region around the curve represents the 95% confidence interval. NDVI was rescaled from 0 to 100. NDVI: Normalized Difference Vegetation Index.
